# Early treatment outcome prediction in metastatic castration-resistant prostate cancer utilizing 3-month tumor growth rate (*g*-rate) based machine learning model

**DOI:** 10.64898/2026.02.26.26346987

**Authors:** Esther C. Ugwueke, Mohamed Azzam, Mengxi Zhou, Benjamin A. Teply, Raymond C. Bergan, Shibiao Wan, Antonio Tito Fojo, Harshraj Leuva, Jieqiong Wang

## Abstract

**Background:** Once the treatment starts, early prediction of treatment benefit and its correlation with overall survival (OS) remains challenging in metastatic castration-resistant prostate cancer (mCRPC). Existing prognostic models require long-term follow-up, limiting their ability to inform timely treatment decisions. To address this gap, we evaluated tumor growth rate (***g***-rate)–based survival models across multiple treatment lines to assess their ability to predict OS and support early clinical decision-making.

**Methods:** We developed GxSurv, a Random Survival Forest (RSF)–based framework that incorporates baseline clinical variables and ***g***-rate calculated from serial on-treatment PSA, to construct line-specific prediction models of OS, a direct measure of treatment outcome. Three variants were developed: G3Surv, using the 3-month ***g***-rate; G6Surv, using the 6-month ***g***-rate; and GfSurv, using the final observed ***g***-rate. Model performance was evaluated using Harrell’s C-index, Uno’s C-index, Integrated Brier Score (IBS), time-dependent area under the curve (tAUC). Model interpretability was assessed using permutation importance to quantify predictor contributions within the GxSurv framework.

**Findings:** The study included 15912 treatment records from 11014 patients with mCPRC across four lines of therapy. We found that incorporation of ***g***-rate consistently improved model performance across all treatment lines, with all GxSurv models outperforming Cox proportional hazards (CoxPH). As the earliest prognostic model, our G3Surv demonstrated strong early predictive performance, with Harrell’s C-index values ranging from 0·700 to 0·746 and tAUC values of 0·766 to 0·822 across all lines, representing 5-8% and 4-5% improvements over CoxPH, respectively. These results indicate that G3Surv accurately predicts individual treatment outcomes at 3 months after treatment initiation. Feature importance analyses consistently identified ***g***-rate as a top predictor, followed by baseline PSA and hemoglobin, with relative variation across treatment lines.

**Interpretation:** Integrating ***g***-rate calculated from on-treatment PSA values enables accurate, line-specific prediction of treatment outcomes in mCRPC, with the 3-month ***g***-rate providing robust early prognostic information to support timely, personalized clinical decision-making.

**Funding:** U.S. National Science Foundation, National Institutes of Health, American Cancer Society.

## 1. Introduction

In the United States, prostate cancer (PC) is the second most commonly diagnosed cancer, with approximately 15·4% of the male population diagnosed with new cases and 5·8% mortality in 2025.^1^ Although five-year relative survival for PC is high in localized disease, it drops markedly to 37·9% amongst patients with distant metastatic disease,^1^ with outcomes further influenced by age, comorbidities, and treatment strategies.^2^ Notably, approximately 8% of patients present with metastatic disease initial diagnosis.^3^ Although many early-stage PC patients initially respond well to therapy, most eventually progress to castration-resistant prostate cancer (CRPC), which remains incurable despite the availability of newer systemic therapies.^4^ In the metastatic castration-resistant prostate cancer (mCRPC) setting, prognosis remains limited despite therapeutic advances, with median overall survival typically ranging from 1.5 to 2.5 years depending on treatment sequence and prior therapies.^5^ Patients who develop mCRPC experience poor and heterogeneous outcomes. Thus, accurate and early prediction of treatment outcomes is essential to guide timely treatment switching, optimize therapy sequencing, and support individualized clinical decision-making.

Numerous survival prediction models have been developed for prostate cancer to support prognostic assessment and clinical decision-making.^6,7^ Traditional approaches using Cox proportional hazards (CoxPH) regression and, more recently, machine learning (ML) methodologies have been applied across various disease states, from localized to metastatic disease.^8–14^ However, the applicability of these models to mCRPC remains limited, as mCRPC represent a unique and clinically complex disease condition marked by androgen deprivation therapy (ADT) resistance,^15^ rapid disease progression, and variability in survival.^16,17^ These clinical and biological characteristics substantially challenge the assumptions underlying conventional prognostic approaches.

Existing prognostic models for mCRPC have primarily relied on CoxPH approaches with heterogeneous baseline clinical variables to predict overall survival (OS).^18–25^ However, these models face four critical limitations: (1) they assume linear covariate effects and proportional hazards over time, assumptions that are frequently violated in mCRPC, where disease biology is highly heterogeneous and evolves rapidly under successive lines of therapy, resulting in only modest predictive performance;^26^ (2) many models are developed within narrowly defined treatment,^20,27,28^ or focus on overall treatment effects, thereby limiting their generalizability across sequential lines of therapy; (3) they do not enable early, response-informed prognostic assessment, as predictions are generally based on baseline data and are not conditioned on early on-treatment tumor response during the initial treatment period;^18–25^ (4) most models do not incorporate dynamic biomarkers like tumor growth rate (***g***-rate) that capture early treatment response, tumor kinetics and has robust inverse correlation with OS.^29–33^ Collectively, these limitations underscore the need for survival prediction models that capture nonlinear biological relationships, evolve to adapt treatment contexts, and integrate early disease dynamics to support clinical decision-making in mCRPC.

To address these limitations, we developed GxSurv, a machine learning framework that predicts individual treatment outcome, as measured by OS probability, for patients with mCRPC across treatment lines. The framework integrates dynamic biomarkers (***g***-rate) with baseline clinical variables to capture both early treatment response and patient disease characteristics.

## 2. Methods

### 2.1 Data Source

Our study utilized data from VA Corporate Data Warehouse (CDW) using VA Informatics and Computing Infrastructure (VINCI). Patients diagnosed with PC were identified using ICD-9/ICD-10 codes and matched with oncology registry records for diagnosis verification.^34^ This cohort study was reviewed and approved by the relevant institutional review board, with a waiver consent. The records included demographics variables, Gleason scores, comorbidities, and treatment-related laboratory values. For each treatment line, we had data on individual treatment start and end dates, baseline and on-treatment serial PSA levels, baseline hemoglobin (HGB), baseline white blood count (WBC), baseline absolute neutrophil count (ANC), and baseline platelets (PLT). Survival time was calculated as the time from treatment initiation to the date of death or last available follow-up, whichever occurred first. This line-specific survival time was used as a direct measure of treatment outcome. All data were anonymized, and dates were converted into intervals before any further analyses.

To measure tumor growth rate, we calculated the ***g***-rate as described in our previous publications.^29–32^ In brief, we use the mathematical model f (t) = exp (−d * t) + exp (g * t) – 1 to measure tumor growth and regression. The total tumor burden f(t) consists of two parallel exponential processes where therapy-resistant cells grow at rate **g/day** and therapy-sensitive cells decrease at rate **d/day** at time (t).^29,32^ This method utilized is a previously validated approach in patients with prostate cancer using clinical trials and VINCI data.^29,31–33^ Tumor growth [***g***] and regression [***d***] were both calculated using on-treatment serial PSA values and the TUMGr package for R (https://cran.r-project.org/web/packages/tumgr/index.html).^35^ We calculated tumor growth rates (***g***-rate) using on-treatment PSA values at three time points: only the first 3 months of PSA, only the first 6 months of PSA, and all available on-treatment PSA to evaluate both short-term treatment effects and extended disease progression.

### 2.2 Data selection

We used the following criteria to select patients and treatment records: (1) mCRPC patients with calculable ***g***-rates, (2) no missing or null data values, and (3) no errors in follow-up data for survival calculation. **Figure 1** shows the data selection process and the number of cohorts in each line of therapy. A final total of 15912 records from 11014 mCRPC patients were included in the study, including 8899 patients in Line 1, 4195 in Line 2, 1899 in Line 3771 in Line 4 and 148 in Line 5 and beyond. Analyses were focused only on participants in the first to fourth treatment lines due to limited patient numbers in later lines of therapy.

**Figure 1:**
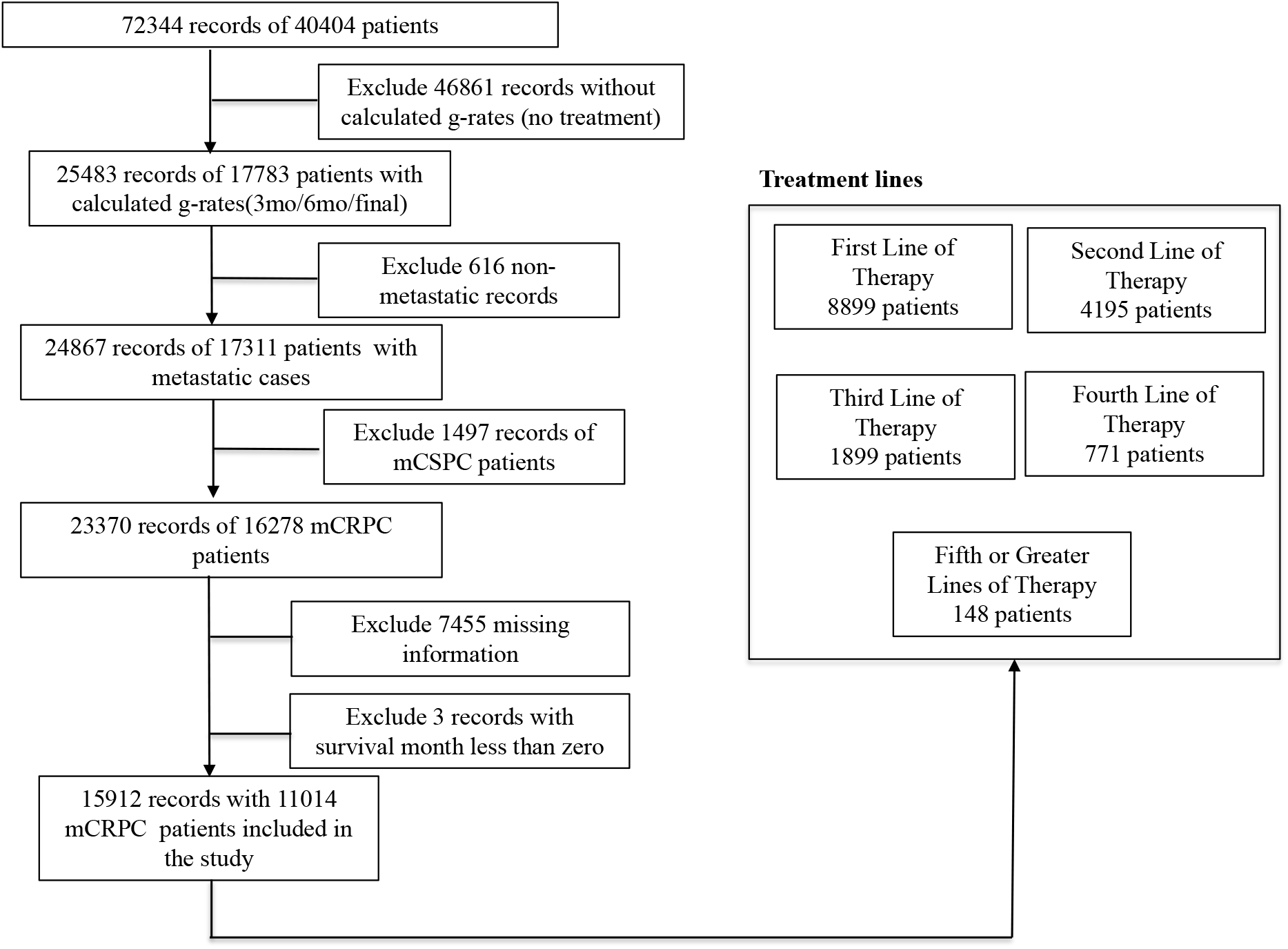
Data selection and cohort construction workflow. Beginning with the full patient dataset, individuals with metastatic castration-resistant prostate cancer (mCRPC) were identified and further categorized into treatment-line subgroups. Due to smaller sample sizes, the fifth to seventh treatment lines were combined into a single category (≥5). The final study cohort reflects the inclusion criteria, exclusion steps, and treatment-line stratification applied to construct the dataset for downstream analyses.

### 2.3 Model development

We developed the survival prediction models with treatment-specific ***g***-rate and the nine most commonly used clinical features obtained at the first visit of treatment start. The nine features includes age, Charlson Comorbidity Index without Cancer (cciwocancer), PSA at diagnosis, Gleason scores, as well as five measures obtained at the start of treatment, i.e. PSA, Hemoglobin (HGB), White Blood Cell (WBC), Platelet (PLT), and Absolute Neutrophil Count (ANC). The dataset was split using stratified sampling for survival status to balance the representation of outcomes between the training (80%) and test (20%) sets for each treatment line. We used the trainset to train the models using the variables without ***g***-rate and with the ***g***-rate measurements at 3 months, 6 months and final, then evaluated the model performance by the independent test set. These models were denoted as GxSurv, where x represents the ***g***-rate assessment window: G0Surv (without ***g***-rate), G3Surv (3-month ***g***-rate), G6Surv (6-month ***g***-rate), and GfSurv (final ***g***-rate). We utilized RSF as our machine learning approach for survival analysis. The RSF approach is an extension of Breiman’s Random Forest (RF) technique, specifically designed for right-censored time-to-event data.^36^

### 2.4 Model performance evaluation

We evaluated the model with independent test set. The evaluation metrics include (1) concordance index (C-index),^37^ which measures a model’s discriminative ability to correctly rank individuals according to their risk of experiencing an event. Among its variants,^37^ Harrel’s C-index depends on the censoring pattern hence simply discards censored pairs in the data to estimate discrimination, while Uno’s C-index,^38^ measure corrects censoring bias using inverse probability of censoring and makes the estimate independent of the censoring distribution; (2) time-dependent area under curve (tAUC),^37^ which evaluates the model’s performance at specific timepoints. A higher tAUC value indicates that the model is significantly proficient in differentiating how well a model can separate subjects who experienced an event from those who do not or rather survive beyond that time; (3) the Integrated Brier Score (IBS), which measures the overall average performance for the prediction model for all observed times.^37^ Besides these, model calibration was assessed by evaluating the calibration slope, which ideally equals 1·0, and by examining the absolute differences between predicted and observed event probabilities. Model performance was further evaluated across different time horizons and compared against a baseline CoxPH model.

### 2.5 Model’s interpretation

To interpret the ML model, we utilized the permutation-based variable importance within the RSF framework. This approach identifies the features that are most important for the model’s predictions by quantifiying their impact on predictive performance. Random Forest variable importance (VIMP), also known as permutation importance, is a widely used interpretability measure in survival modeling.^36,39^ The VIMP technique reveals the extent to which each variable influences the survival outcome and assesses the degree to which the prediction accuracy dimimshes when the relationship between the variable and survival is randomly disrupted. This feature importance evaluation provides clinical interpretation of key prognostic indictaors.^36,40^

## 3. Results

### 3.1 Cohort Characteristics

The first-line mCRPC cohort consisted of 8899 patients with a median age of 72·7 years and 72·8 years in the training set and test sets, respectively. Median survival times in the training and test sets were 20·6 months (IQR 10·4–36·8) and 20·8 months (IQR 10·6–36·8), respectively. The second-line mCRPC cohort consisted of 4195 patients with a median age of 72·9 years and median survival times of 13·9 months (IQR 7·2–25·2) in the training set aÏnd 72·2 years and a median survival of 14·4 months (IQR 7·3–26·2) in the test set. In the third line of therapy, a total of 1899 patients were split into training and test sets, with median ages of 71·6 and 71·5 years, respectively, and median survival times of 11·6 months (IQR 6·2–20·1) and 11·0 months (IQR 6·1–19·5), respectively. The fourth line of therapy with 771 patients had a median survival time of 8·7 months (IQR 4·7–15·7) in the training set and 8·5 months (IQR 5·5–14·2) in the test set, with a median age of 71·1 and 72·2 years in the training and test sets, respectively. The observed median survival times differed among the four different lines of therapy. For additional details on patient characteristics, refer to **Supplementary Tables 1-4**.

### 3.2 Model performance with *g*-rate at different time points

We evaluated the performance of line-specific models on independent test sets, with survival prediction results summarized in **Table 1** and **Figure 2**. Across all treatment lines, incorporation of ***g***-rate substantially improved the discriminative performance of the models, with relative increases of approximately 5-12% in Harrell’s C-index and 5-7% in tAUC compared with models that excluded ***g***-rate. In the first treatment line, the C-index increased from 0·712 without the ***g***-rate to 0·761 with the final ***g***-rate. The second line showed a similar trend, moving from 0·688 to 0·754. In the third line, C-index increased from 0·647 to 0·724. The fourth treatment line followed suit rising from 0·625 to 0·701 after the final ***g***-rate was added. Overall, models incorporating the final ***g***-rate achieved a robust performance in predicting survival across all treatment lines. This improvement likely reflects the ability of the final ***g***-rate to capture the stable, treatment-resistant tumor growth dynamics, thereby providing complementary prognostic information beyond baseline clinical variables.

**Table 1.** Comparison of survival prediction model performance across four treatment lines using GxSurv models. Censorship (%) indicates the proportion of participants in each treatment line. Values of evaluation metrics represent point estimates with 95% confidence intervals in parentheses. G0Surv: model without tumor growth rate; G3Surv: model with 3-month ***g***-rate; G6Surv: model with 6-month ***g***-rate; and GfSurv: model with final-observed ***g***-rate. IBS: integrated brier score, tAUC: time dependent area under curve. CI: confidence interval.

| Treatment Line | Censorship (%) | Models | Harrel's C-index (95% CI) ↑ | Uno's C-index (95% CI) ↑ | IBS (95% CI) ↓ | tAUC (95% CI) ↑ |
| --- | --- | --- | --- | --- | --- | --- |
| First line | 23·5 | G0Surv | 0·712<br>(0·697-0·726) | 0·698<br>(0·684-0·711) | 0·081<br>(0·071-0·092) | 0·780<br>(0·762-0·798) |
|  |  | G3Surv | 0·746<br>(0·733-0·759) | 0·729<br>(0·716-0·742) | 0·077<br>(0·068-0·087) | 0·822<br>(0·806-0·839) |
|  |  | G6Surv | 0·754<br>(0·741-0·766) | 0·736<br>(0·723-0·749) | 0·076<br>(0·067-0·087) | 0·831<br>(0·815-0·847) |
|  |  | GfSurv | 0·761<br>(0·748-0·775) | 0·745<br>(0·732-0·759) | 0·074<br>(0·066-0·085) | 0·836<br>(0·820-0·852) |
| Second line | 16·0 | G0Surv | 0·688<br>(0·667-0·706) | 0·678<br>(0·658-0·696) | 0·100<br>(0·090-0·112) | 0·760<br>(0·732-0·784) |
|  |  | G3Surv | 0·737<br>(0·719-0·755) | 0·726<br>(0·710-0·744) | 0·091<br>(0·082-0·102) | 0·818<br>(0·797-0·840) |
|  |  | G6Surv | 0·743<br>(0·726-0·761) | 0·733<br>(0·716-0·750) | 0·088<br>(0·079-0·100) | 0·825<br>(0·804-0·845) |
|  |  | GfSurv | 0·754<br>(0·737-0·772) | 0·744<br>(0·727-0·761) | 0·085<br>(0·076-0·095) | 0·835<br>(0·814-0·855) |
| Third line | 13·7 | G0Surv | 0·647<br>(0·612-0·680) | 0·643<br>(0·607-0·675) | 0·083<br>(0·070-0·106) | 0·707<br>(0·660-0·751) |
|  |  | G3Surv | 0·704<br>(0·675-0·731) | 0·697<br>(0·668-0·725) | 0·076<br>(0·063-0·097) | 0·786<br>(0·750-0·819) |
|  |  | G6Surv | 0·717<br>(0·688-0·745) | 0·709<br>(0·680-0·737) | 0·076<br>(0·064-0·098) | 0·801<br>(0·765-0·835) |
|  |  | GfSurv | 0·724<br>(0·696-0·750) | 0·716<br>(0·688-0·743) | 0·075<br>(0·063-0·096) | 0·809<br>(0·774-0·842) |
| Fourth line | 13·6 | G0Surv | 0·625<br>(0·569-0·680) | 0·621<br>(0·566-0·676) | 0·105<br>(0·085-0·132) | 0·664<br>(0·590-0·741) |
|  |  | G3Surv | 0·700<br>(0·647-0·750) | 0·694<br>(0·643-0·742) | 0·096<br>(0·077-0·120) | 0·766<br>(0·700-0·830) |
|  |  | G6Surv | 0·698<br>(0·649-0·745) | 0·694<br>(0·646-0·739) | 0·094<br>(0·076-0·118) | 0·763<br>(0·696-0·825) |
|  |  | GfSurv | 0·701<br>(0·651-0·749) | 0·697<br>(0·649-0·744) | 0·093<br>(0·075-0·117) | 0·766<br>(0·700-0·828) |

**Figure 2:**
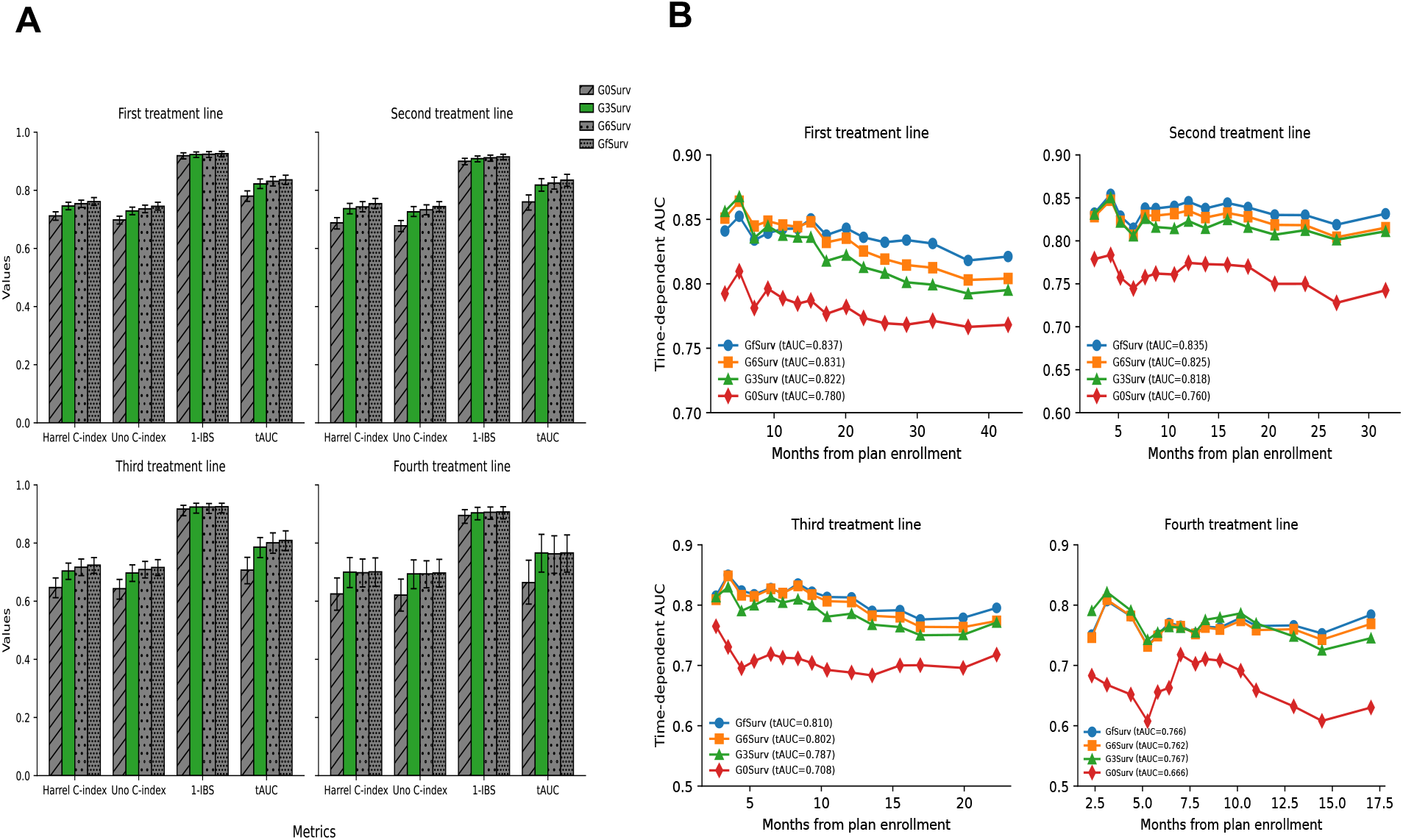
Performance of survival prediction models across four treatment lines. (A) Bar plots comparing predictive performance across four treatment lines for models without tumor growth rate (G0Surv) and models incorporating ***g***-rate at 3 months (G3Surv), 6 months (G6Surv), and the final time point (GfSurv). All lines show models incorporating ***g***-rate demonstrate superior discrimination and lower prediction error compared to baseline. (B) Time-dependent AUC curves of different survival prediction models across four treatment lines. G-rate: tumor growth rate, IBS: integrated brier score, tAUC: time dependent area under curve.

Notably, the G3Surv model showed consistently strong predictive performance across all treatment lines, with performance comparable to that of G6Surv and GfSurv. The G3Surv achieved a C-index and tAUC values of 0·746 and 0·822 in the first line, 0·737 and 0·818 in the second line, 0·704 and 0·786 in the third line, and 0·700 and 0·766 in the fourth line, demonstrating consistently robust predictive ability in early treatment across all lines (**Table 1**). We focused on evaluating G3Surv performance because it could help make early treatment decisions in clinics once it is prospectively validated.

### 3.3 Comparison with other models

**Figure 3** presents a comparison of G3Surv versus CoxPH across treatment lines. G3Surv demonstrated superior predictive performance across all treatment lines, with the most pronounced improvements in earlier therapy. For the first and second treatment lines, G3Surv, as part of the GxSurv framework, showed substantial advantages: C-indices were 5·07% and 4·84% higher for the G3Surv models compared to CoxPH models (0·746 vs 0·710 and 0·737 vs 0·703, respectively), and integrated Brier scores were dramatically reduced by 52·8% and 45·8% (0·077 vs 0·163 and 0·091 vs 0·168, respectively), indicating markedly improved prediction accuracy. Consistently, tAUC values for G3Surv (0·822–0·818) corresponded to approximately 5·0% and 4·5% relative improvements, respectively, compared with the CoxPH model (0·783), further corroborating the enhanced performance. While both models showed reduced performance in later treatment lines due to increased heterogeneity and censoring, G3Surv maintained a consistent predictive advantage with higher C-indices and persistently lower IBS scores. This superior performance was replicated across G6Surv and GfSurv models (**Supplementary Table 6**). These findings demonstrate that our GxSurv models are substantially superior to Cox regression for OS prediction in mCRPC, particularly in early treatment decisions.

**Figure 3:**
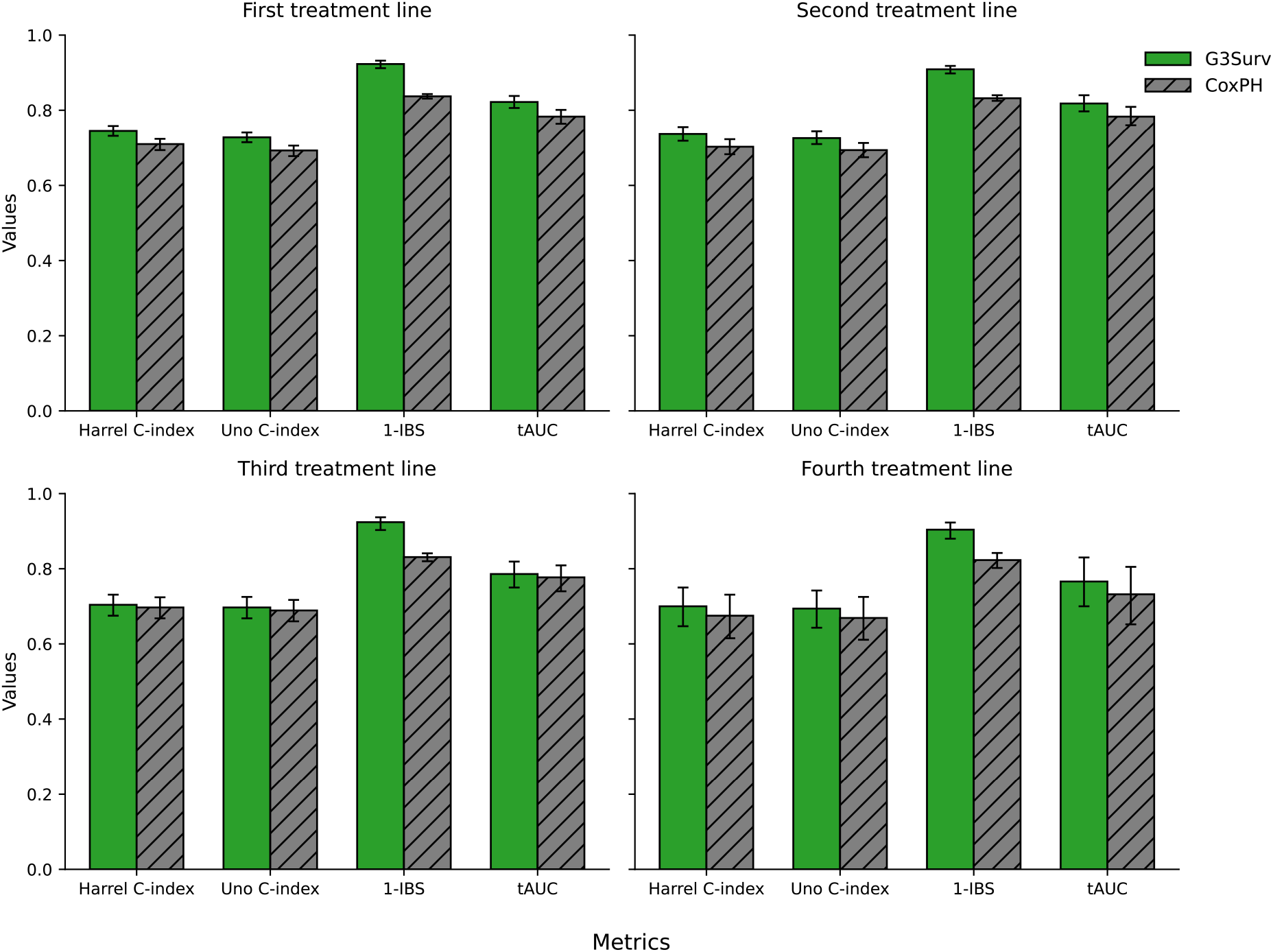
Performance comparision of survival prediction between G3Surv and CoxPH model. Bar graphs illustrate Harrell’s C-index, Uno’s C-index, 1-IBS, and tAUC for the G3Surv (green) and CoxPH (gray) models throughout the first to fourth treatment lines. Our G3Surv model (green) exhibits superior discrimination and reduced prediction error compared to the CoxPH model across all treatment lines. Error bars denote 95% confidence intervals. IBS: Integrated brier score, tAUC:time dependent area under curve.

### 3.4 Survival curve obtained by the G3Surv model

**Figure 4** illustrates the predicted survival curves, i.e., treatment outcomes, of twelve representative patients across four treatment lines via G3Surv models. Patients with faster ***g***-rates showed much earlier event times and drops in survival probability, whereas patients with slower ***g***-rate experienced gradual declines and later event times. All observed events in the first treatment line occurred between 10 and 40 months. In the second treatment line, the curves begin to cluster more tightly, with most events occurring between 10 and 25 months, reflecting shorter survival and reduced variability as patients move into later therapy. In the third treatment line, survival drops more steeply, and nearly all events occur before 20 months. The fourth treatment line shows the most compressed curves, with rapid early declines and event times predominantly within the first 25 months. Together, the four panels illustrate the progressive shortening and tightening of survival trajectories across treatment lines and visually reinforce the strong association between ***g***-rate and survival.

**Figure 4:**
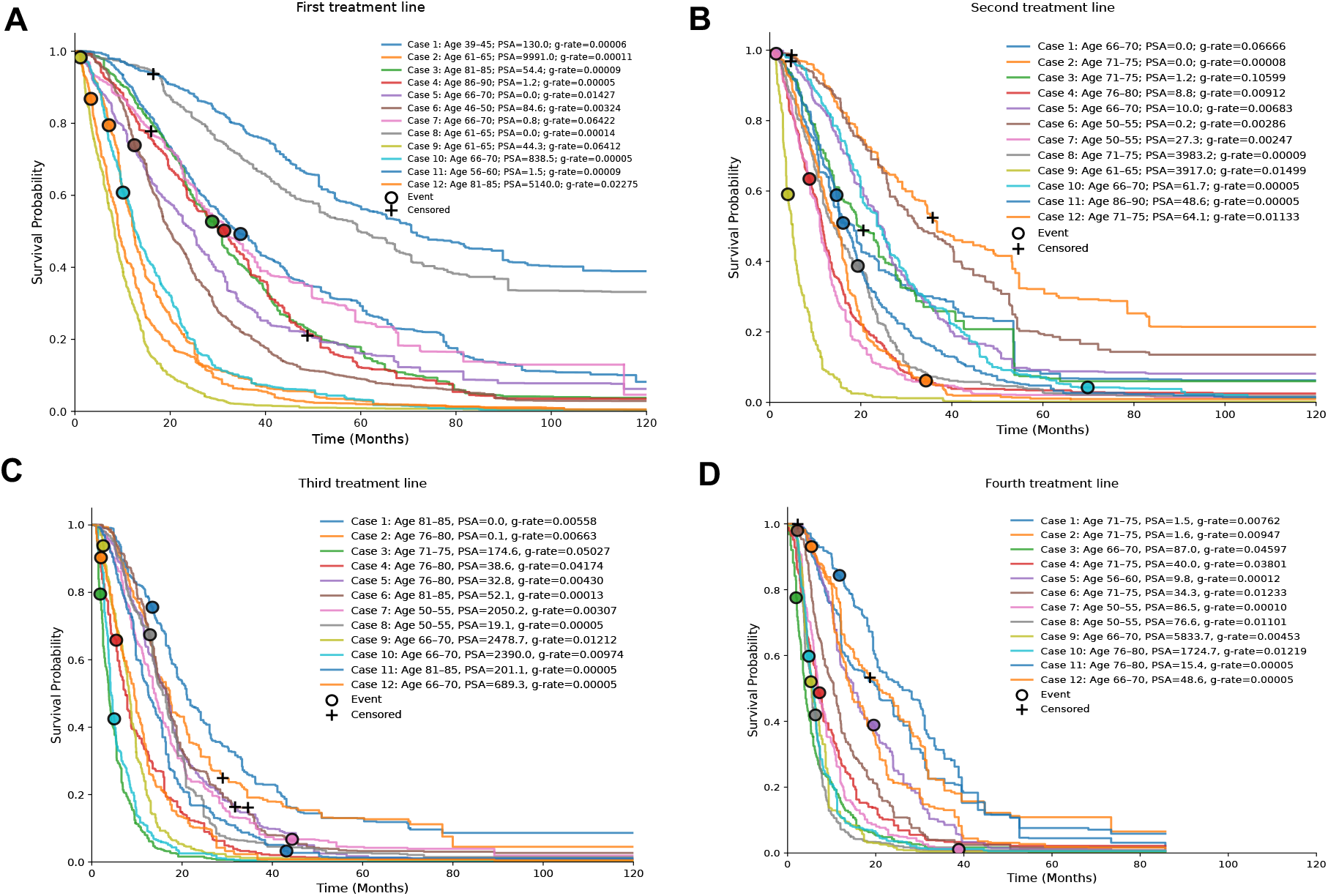
Survival curves for 12 representative individual mCRPC patients predicted by G3Surv prediction models. Panels A–D correspond to Treatment Lines 1-4. Representative patients were selected to represent the full spectrum of clinical (age, baseline PSA) and tumor-kinetic (g-rate) variability. Heterogeneous survival trajectories reflect substantial inter-patient differences in treatment response and disease progression. ○: event. +: censored.

### 3.5 Interpretation of the survival prediction model

**Figure 5** illustrates the importance of variables in predicting OS across treatment lines using G3Surv. The ***g***-rate was consistently the strongest predictor of survival across all lines of therapy. Other highly ranked predictors include PSA and HGB at treatment initiation. The ranking of the PSA and HGB in the first and second line of treatment remained consistent, while HGB surpassed PSA in subsequent treatment lines. Conventional clinical indicators, including Gleason score, age, ANC, WBC, and PLT, provided relatively little predictive value, whereas the comorbidity index and PSA at diagnosis offered minimal contributions across all models. Similar variable rankings were observed for G6Surv and GfSurv models (**Supplementary Figure 4** and **Supplementary Figure 5**). We further evaluated the contribution of the three least important measures (PSA at diagnosis,WBC, cciwocancer) by excluding them from the GxSurv models. Their removal had a negligible effect on OS prediction compared with the full ten-feature models (**Supplementary Table 5)**.

**Figure 5:**
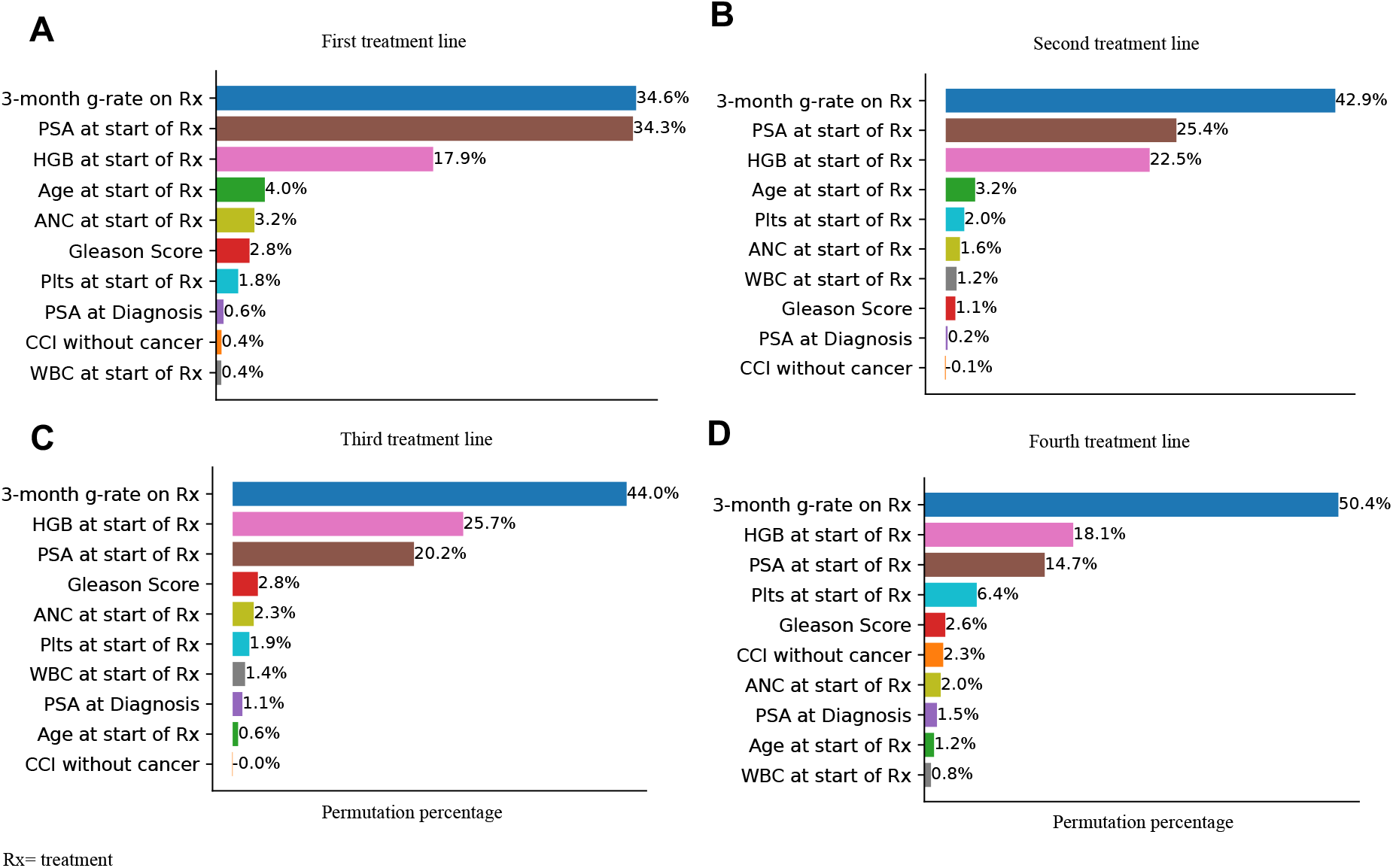
Feature importance of G3Surv model across treatment lines. Panels A–D correspond to Treatment Lines 1-4. The g-rate was the most influential predictor of overall survival, with its importance progressively increasing from the first through the fourth treatment line.

## 4. Discussion

In this study, using data from 15912 treatment records, we developed novel GxSurv models via ***g***-rate and most-commonly used clinical variables to predict treatment outcome, quantified by OS probability, in patients with mCRPC. Our G3Surv model can accurately predict the personalized OS as early as 3 months from the start of therapy across Line1-4. The top 3 clinical variables in the ML model are ***g***-rate, starting PSA and starting HGB. To our knowledge, our study is the first to develop an individualized OS prediction model for specific treatment lines for patients with mCRPC, demonstrating remarkable progress toward more precise predictions. Our G3Surv model used only the on-treatment PSA during the first 3 months to calculate ***g***-rate, along with 9 clinical variables, providing physicians worldwide with a practical tool to guide early decision-making during treatment.

Early prognostic assessment is critical for guiding timely treatment decisions and patient counseling in mCRPC. We therefore prioritize the G3Surv, which enables the prediction of treatment outcome at 3 months, a clinically actionable timepoint when treatment modifications can meaningfully impact outcomes. Our approach is supported by prior studies demonstrating that early PSA dynamics within the first twelve weeks are strongly associated with overall survival.^41,42^ Compared with the G3Surv model, our GfSurv models (using all on-treatment PSA to calculate ***g***-rate) showed marginally improved performance metrics; these gains did not translate into clinically meaningful differences in survival predictions. Importantly, the actionability of early (3-month) treatment outcome prediction via G3Surv provides substantially greater clinical value than the minor improvements in prognostic performance metrics (C-index, IBS, tAUC) offered by delayed assessment with later ***g***-rate models.

Our GxSurv models demonstrated superior predictive accuracy compared to CoxPH regression across all treatment lines, consistently achieving higher C-indices and tAUC, along with lower IBS (**Supplementary Table 6 and Supplementary Table 7**). Notably, these improvements were most pronounced in early treatment lines, where accurate prognostic stratification carries the greatest clinical consequence. GxSurv achieves superior performance through its use of RSF, which uniquely captures non-linear relationships and complex interactions among clinical and laboratory variables that CoxPH’s proportional hazards assumption cannot adequately represent. The reproducibility of these performance enhancements across various g-rate specifications and evaluation criteria supports the stability of RSF, rather than indicating a model-specific phenomenon. Our findings support previous research highlighting the constraints of proportional hazards assumptions in advanced cancer cohorts and the advantage of more flexible survival modeling approaches in capturing non-linear effects and higher-order interactions.^43,44^

Compared with previously published survival prediction models in mCRPC,^21,22^ GxSurv demonstrates consistently improved prognostic performance. Prior survival prediction studies in mCRPC have several limitations: they typically did not stratify analyses by treatment line,^45–47^ relied on relatively homogeneous patient cohorts, and employed static or summary PSA measures ^22,46^ that inadequately capture longitudinal tumor dynamics driven by effects of treatment.^46,47^In contrast, our GxSurv models were specifically developed for heterogeneous real-world mCRPC populations and evaluated across distinct treatment lines. Across all evaluated treatment lines, our models consistently demonstrated superior performance compared with previously published models (**Supplementary Table 8**).^21,22^ Consistent with our previously validated studies,^29,32^ the current findings confirm that ***g***-rate provides more informative prognostic information than conventional PSA and serves as an excellent biomarker for OS.

We demonstrated the seven most significant predictors of treatment outcome. Utilizing an initial set of 10 variables, GxSurv models exhibited robust discriminative efficacy across all therapeutic lines (**Table 1 and Figure 2A**). A reduced seven-variable model (**Supplementary Table 5**) based on permutation feature importance retaining age, Gleason score, PSA, HGB, PLT and ANC at baseline, and the ***g***-rate showed the stability of these predictors and enhanced clinical usability. This simplified model exhibited performance equivalent to that of the full model across all treatment lines (**Supplementary Table 5)**. This suggests that a small, clinically intuitive set of features is sufficient to capture most of the information about survival, supporting the idea that clinicians can focus on the top seven features to predict treatment outcome.

The top 3 clinical measurements identified by our GxSurv models across all treatment lines are ***g***-rate, baseline PSA, and baseline HGB (**Figure 5**). ***G***-rate consistently emerges as the variable most strongly associated with overall survival. G-rate compresses complex longitudinal PSA dynamics, baseline burden, regression magnitude, nadir behavior, and subsequent regrowth into a single parameter that directly reflects tumour biology and treatment resistance. As a result, it captures prognostic information not obtainable from static clinical variables, making it the most informative predictor of treatment outcome in metastatic disease. Accordingly, these findings support our previous finding that ***g***-rate serves as a better survival predictor than PSA doubling time and all other PSA-based summary values.^30,31^

PSA ranks 2^nd^ and HGB ranks 3^rd^ in the first two lines whereas HGB ranks 2^nd^ and PSA ranks 3^rd^ in the Line 3 & 4 (**Figure 5)**. From Line 3 onward, most patients have received the chemotherapy and/or radiopharmaceutical treatments. This shift likely reflects cumulative hematologic effects from prior therapies in heavily pretreated mCRPC patients, including chemotherapy and radiopharmaceutical treatments, which can suppress red blood cell production and contribute to anemia.^27^ The results indicate that HGB monitoring needs to become a priority for risk assessment in treatment line-specific evaluations, especially for patients who receive chemotherapy and radiopharmaceuticals. Physicians should consider anemia an independent risk factor affecting patient outcomes, rather than a side effect that requires supportive care.

While our study contributes to early treatment decisions by predicting individual patients’ overall survival at 3 months, several limitations should be considered. First, the lack of an external validation cohort impacts our ability to generalize our findings. Also, there is a clear trend of varied censoring rates across the treatment lines in the test sets, ranging from 23·5% in the first line to 13·5% in the fourth line, which may affect predictive accuracy and lead to a decline in later treatment lines. Our model was developed using real-world veterans data and not randomiazed clinical trial data recorded at a specific time point,s and that can introduce selection bias; however, it also serves as a proof of its applicability in the real wold use case where timing of lab collection is highly varied. While some previously identified prognostic factors were not used in this study, incorporating these variables with ***g***-rate, along with external prospective cohort validation, will further enhance the accuracy of our models.

## 5. Conclusion

This study developed GxSurv, a series of machine learning models that predict individualized treatment outcomes at different time points after treatment initiation for patients with mCRPC. Our G3Surv model accurately predicted overall survival at the individual-patient level across 1–4 treatment lines in mCRPC using data from only the first 3 months of treatment. The model’s interpretability identified key prognostic variables to be ***g***-rate, PSA, HGB, ANC, and PLT at the start of treatment; Gleason score; and age. Future efforts should focus on assessing the addition of other commonly collected variables for treatment and on external validation to ensure broad clinical relevance.

## Data Availability

The US Veterans Health Administration prohibits sharing data unless there is a signed data access agreement with the VA research team requesting it and it is approved by the IRB.

## Research in context

**Evidence before this study**

Most previous studies about mCRPC prognostic modeling have concentrated on long-term survival results but have not examined effect of treatment on tumor dynamics. These survival models exist for extended follow-up periods, which reduces their value for making individualized early treatment choices. The identification of patients who face elevated risks during their first three months of treatment has not received sufficient research attention for personalized treatment changes. We searched PubMed database from inception to August 30,2025 using the terms “prostate cancer” or “metastatic prostate cancer” or “mCRPC” AND “survival prediction” AND “Machine learning”. We reviewed several papers based on our search and found studies that utilized baseline variables, but none utilized tumor growth rate (***g***-rate) or predicted survival at 3months timepoint.

**Added value of this study**

In patients with mCRPC, we developed GxSurv models including variants without ***g***-rate, and g-rate calculated using serial on treatment PSA available at first 3-month, 6-month and treatment end across first to fourth treatment lines. Our study showed that incorporation of early ***g***-rate measurements enables accurate and clinically meaningful survival prediction throughout sequential therapies. We demonstrate that ***g***-rate functions as a strong dynamic biomarker that complements and, in many settings, outperforms traditional baseline predictors by allowing real-time assessment of disease aggressiveness. In addition, we introduce treatment-line–specific predictive models that explicitly account for the evolving clinical context across sequential therapies rather than assuming uniform prognostic behavior throughout the disease course. Finally, we identify key clinical indicators consistently associated with survival across treatment lines thereby providing evidence-based priority factors that can inform patient monitoring, risk stratification, and treatment optimization in routine clinical practice.

**Implications of all the available evidence**

Current findings indicate that early ***g***-rate measurement, particularly at 3 months after treatment initiation, provides a practical and informative prognostic signal for survival prediction in patients with metastatic castration-resistant prostate cancer, enabling early risk stratification and providing more timely therapeutic decision-making insight whether to continue the current treatment or consider to switch to another option. While predictive performance may attenuate in later treatment lines as patient populations become more clinically homogeneous and outcomes worsen, integration of complementary data sources such as molecular, imaging, or additional longitudinal biomarkers has the potential to enhance model discrimination and maintain prognostic utility. Overall, ***g***-rate–based modeling represents a flexible and scalable framework for individualized survival prediction that can inform adaptive treatment planning and improve the design and efficiency of future clinical studies.

## Acknowledgements

Research reported in this publication was supported by the U.S. National Science Foundation under Award Number 2500836, the Office Of The Director, National Institutes Of Health of the National Institutes of Health under Award Number R03OD038391, and by the National Cancer Institute of the National Institutes of Health under Award Number P30CA036727. This work was supported by the American Cancer Society under award number IRG-22-146-07-IRG, and by the Buffett Cancer Center, which is supported by the National Cancer Institute under award number CA036727. This study was in part financially supported by the Child Health Research Institute at UNMC/Children’s Nebraska. This work was also partially supported by the University of Nebraska Collaboration Initiative Grant from the Nebraska Research Initiative (NRI). The content is solely the responsibility of the authors and does not necessarily represent the official views from the funding organizations.

