## Supplementary Material for "Early treatment outcome prediction in metastatic castration-resistant prostate cancer utilizing 3-month tumor growth rate (*g*-rate) based machine learning model"

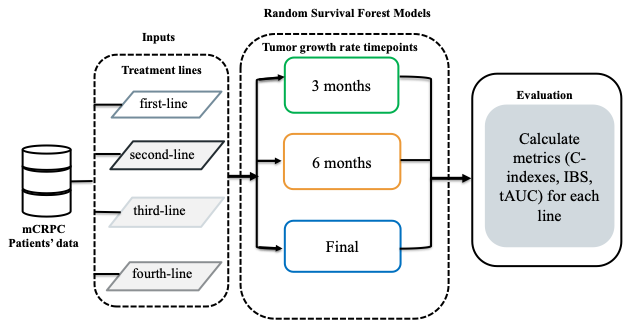

**Supplementary Figure 1: Framework of the machine learning pipeline for individual treatment outcome prediction*.*** A line-specific treatment outcomes workflow for mCRPC utilizing baseline data and tumor growth rates at 3-month, 6-month, and final timepoints as inputs for GxSurv across first- to fourth-line treatments.

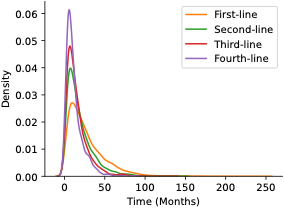

**Supplementary Figure 2: Distribution of survival times of Treatment Line 1-4.**

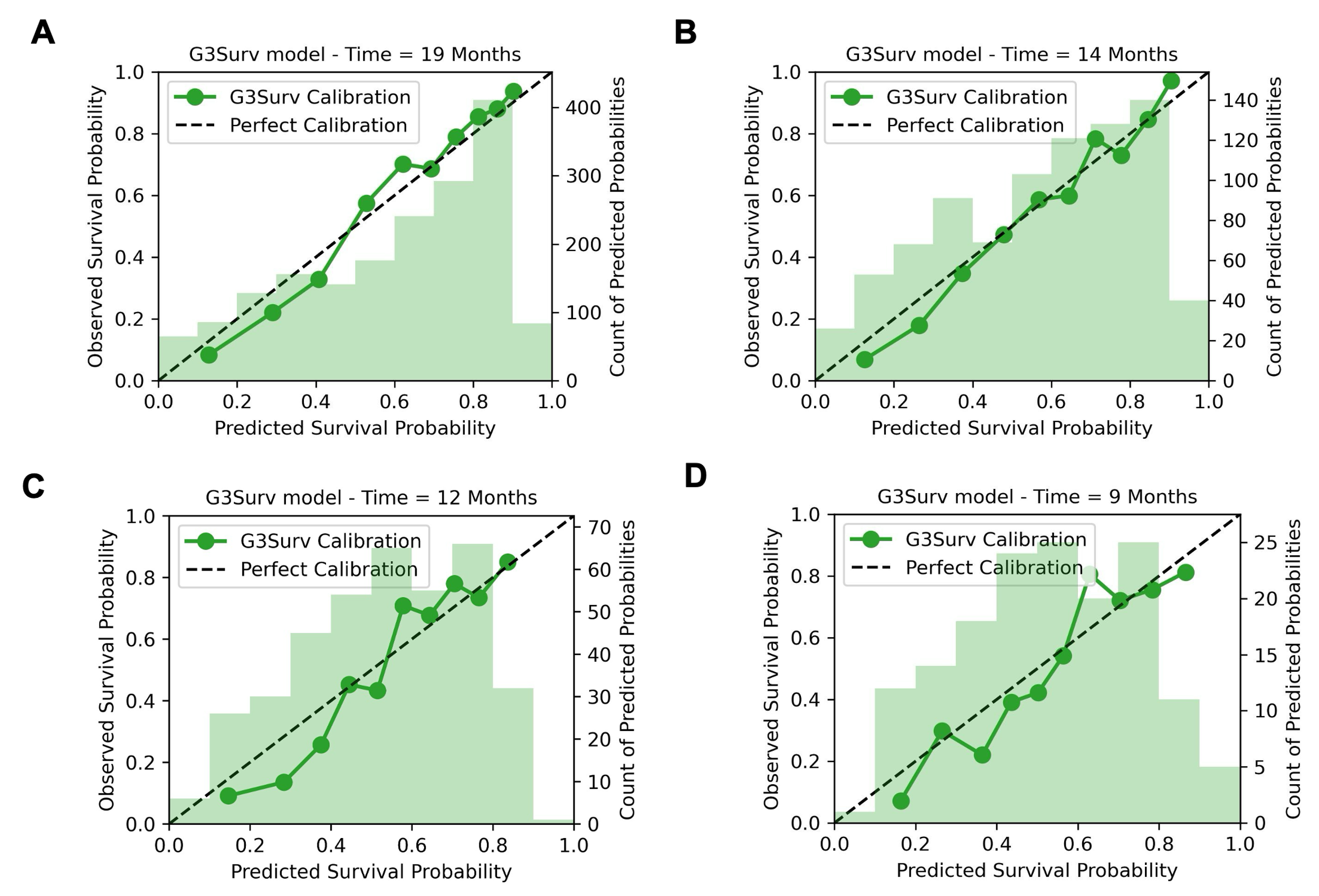

**Supplementary Figure 3**: **Calibration performance of the G3Surv model across treatment lines**. Calibration plots for the GxSurv model in (A) Line 1, (B) Line 2, (C) Line 3, and (D) Line 4. Time = xx months denotes the median survival time for each treatment line, at which calibration was evaluated.

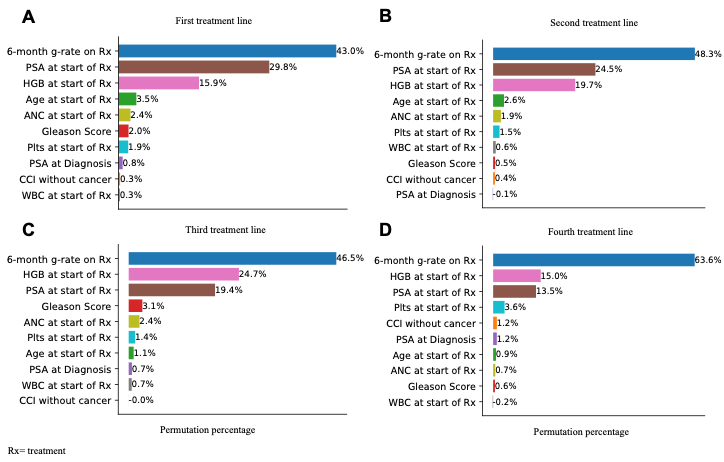

**Supplementary Figure 4: Feature importance of G6Surv model across treatment lines.** Panels A–D correspond to Treament Lines 1-4. The g-rate was the most influential predictor of treatment response, with its importance progressively increasing from the first through the fourth treatment line.

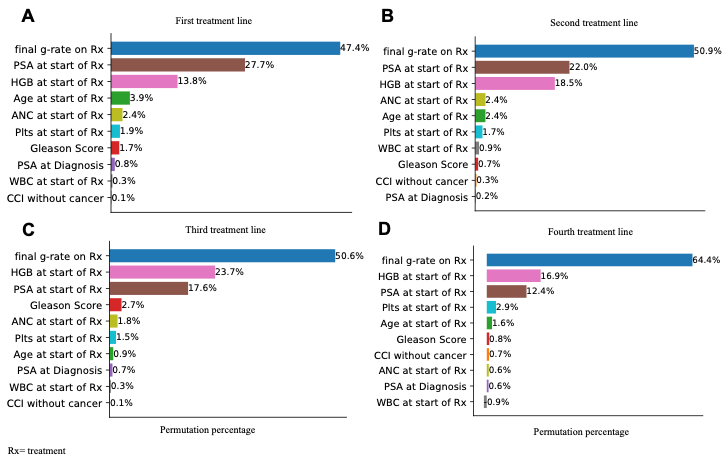

**Supplementary Figure 5: Feature importance of GfSurv model across treatment lines.** Panels A–D correspond to Treament Lines 1-4. The g-rate was the most influential predictor of treatment response, with its importance progressively increasing from the first through the fourth treatment line.

**Supplementary Table 1: Baseline Characteristics between training and test set in the first line of treatment**

|  | **Overall (n=8899)** | **Training (n=7119)** | **Test (n=1780)** | **p_value** |
| --- | --- | --- | --- | --- |
| Age (median [Q1-Q3]) | 72·7 [67·2-79·3] | 72·7 [67·1-79·3] | 72·8 [67·5-79·5] | 0·354 |
| Race (n, %) |  |  |  | 0·462 |
| American Indian or Alaska Native | 56 (0·6%) | 43 (0·6%) | 13 (0·7%) |  |
| Asian | 28 (0·3%) | 22 (0·3%) | 6 (0·3%) |  |
| Black | 2359 (26·5%) | 1917 (26·9%) | 442 (24.8%) |  |
| Native Hawaiian or Other Pacific Islander | 54 (0·6%) | 42 (0·6%) | 12 (0·7%) |  |
| White | 6402 (71.9%) | 5095 (71·6%) | 1307 (73· 4%) |  |
| Charles Comorbidity Index without cancer (n, %) |  |  |  | 0·015 |
| 0 | 1013 (11·4%) | 805 (11·3%) | 208 (11·7%) |  |
| 1 | 1430 (16·1%) | 1131 (15.9%) | 299 (16·8%) |  |
| 2 | 1596 (17·9%) | 1269 (17·8%) | 327 (18·4%) |  |
| 3 | 1494 (16·8%) | 1215 (17·1%) | 279 (15·7%) |  |
| 4 | 1138 (12· 8%) | 914 (12· 8%) | 224 (12· 6%) |  |
| 5 | 822 (9·2%) | 671 (9·4%) | 151 (8·5%) |  |
| 6 | 613 (6·9%) | 490 (6·9%) | 123 (6·9%) |  |
| 7 | 388 (4·4%) | 306 (4·3%) | 82 (4·6%) |  |
| 8 | 209 (2· 3%) | 164 (2· 3%) | 45 (2· 5%) |  |
| 9 | 109 (1·2%) | 90 (1·3%) | 19 (1·1%) |  |
| 10 | 43 (0·5%) | 37 (0·5%) | 6 (0·3%) |  |
| 11 | 21 (0·2%) | 15 (0·2%) | 6 (0·3%) |  |
| 12 | 12 (0·1%) | 5 (0·1%) | 7 (0·4%) |  |
| 13 | 7 (0·1%) | 6 (0·1%) | 1 (0·1%) |  |
| 14 | 2 (0·0%) | 0 (0·0%) | 2 (0·1%) |  |
| 16 | 1 (0·0%) | 1 (0·0%) | 0 (0·0%) |  |
| 17 | 1 (0·0%) | 0 (0·0%) | 1 (0·1%) |  |
| Rurality (n, %) |  |  |  | 0·651 |
| Rural | 2688 (30·2%) | 2142 (30·1%) | 546 (30·7%) |  |
| Urban | 6211 (69·8%) | 4977 (69·9%) | 1234 (69·3%) |  |
| Gleason Score (n, %) |  |  |  | 0·789 |
| 4·0 | 73 (0·8%) | 57 (0·8%) | 16 (0·9%) |  |
| 5·0 | 118 (1·3%) | 89 (1·3%) | 29 (1·6%) |  |
| 5·5 | 49 (0·6%) | 38 (0·5%) | 11 (0·6%) |  |
| 6·0 | 1030 (11·6%) | 825 (11·6%) | 205 (11·5%) |  |
| 6·5 | 15 (0·2%) | 13 (0·2%) | 2 (0·1%) |  |
| 7·0 | 1642 (18·5%) | 1306 (18·3%) | 336 (18·9%) |  |
| 7·5 | 1014 (11·4%) | 827 (11·6%) | 187 (10·5%) |  |
| 8·0 | 1683 (18·9%) | 1336 (18·8%) | 347 (19·5%) |  |
| 8·5 | 75 (0·8%) | 58 (0·8%) | 17 (1·0%) |  |
| 9·0 | 2016 (22· 7%) | 1604 (22· 5%) | 412 (23· 1%) |  |
| 9·5 | 673 (7·6%) | 546 (7·7%) | 127 (7·1%) |  |
| 10·0 | 511 (5·7%) | 420 (5·9%) | 91 (5·1%) |  |
| PSA at Diagnosis (median [Q1-Q3]) | 5·9 [0·8-22· 5] | 5·9 [0·8-22· 7] | 5·8 [0·8-21·7] | 0·383 |
| PSA at start of treatment (median [Q1-Q3]) | 27·9 [7·5-102· 5] | 27·7 [7·5-100·6] | 28·5 [7·4-109·6] | 0·615 |
| g-rate (median [Q1-Q3]) | -7·1 [-9·0--5·4] | -7·0 [-9·1--5·4] | -7·1 [-9·0--5·4] | 0·806 |
| g-rate at 3months (median [Q1-Q3]) | -8·9 [-9·3--5·3] | -8·9 [-9·3--5·3] | -9·0 [-9·3--5·3] | 0·226 |
| g-rate at 6months (median [Q1-Q3]) | -7·2 [-9·2--5·4] | -7·1 [-9·2--5·4] | -7·4 [-9·2--5·4] | 0·368 |
| HGB at start of treatment (median [Q1-Q3]) | 12· 5 [11·2-13· 5] | 12· 4 [11·2-13· 5] | 12· 5 [11·2-13· 6] | 0·417 |
| WBC at start of treatment (median [Q1-Q3]) | 6·9 [5·4-8·7] | 6·9 [5·4-8·7] | 6·9 [5·4-8·8] | 0·427 |
| ANC at start of treatment (median [Q1-Q3]) | 4·5 [3· 3-6·2] | 4·6 [3· 3-6·2] | 4·5 [3· 3-6·1] | 0·932 |
| PLT at start of treatment (median [Q1-Q3]) | 226·0 [184·0-276·0] | 226·0 [184·0-276·0] | 226·0 [185·0-279·0] | 0·519 |
| status (n, %) |  |  |  | 1·000 |
| 0·0 | 2089 (23· 5%) | 1671 (23· 5%) | 418 (23· 5%) |  |
| 1·0 | 6810 (76·5%) | 5448 (76·5%) | 1362 (76·5%) |  |
| time (median [Q1-Q3]) | 20·7 [10·5-36·8] | 20·6 [10·4-36·8] | 20·8 [10·6-36·8] | 0·905 |
| *p-values calculated using Chi-square test for categorical variables and Mann-Whitney U test for continuous variables.  Abbreviations: PSA, prostate-specific antigen (PSA); HGB, Hemoglobin; WBC, White Blood Cell Count; ANC, Absolute Neutrophil Count; PLT, Platelet. | | | | |

**Supplementary Table 2: Baseline Characteristics between training and test set in the second line of treatment**

|  | **Overall (n=4195)** | **Training (n=3356)** | **Test (n=839)** | **p_value** |
| --- | --- | --- | --- | --- |
| Age (median [Q1-Q3]) | 72·8 [67·7-79·1] | 72·9 [67·8-79·1] | 72·2 [67·0-79·1] | 0·069 |
| Race (n, %) |  |  |  | 0·286 |
| American Indian or Alaska Native | 23 (0·5%) | 20 (0·6%) | 3 (0·4%) |  |
| Asian | 12 (0·3%) | 7 (0·2%) | 5 (0·6%) |  |
| Black | 1124 (26·8%) | 891 (26·5%) | 233 (27·8%) |  |
| Native Hawaiian or Other Pacific Islander | 30 (0·7%) | 23 (0·7%) | 7 (0·8%) |  |
| White | 3006 (71·7%) | 2415 (72· 0%) | 591 (70·4%) |  |
| Charles Comorbidity Index without cancer (n, %) |  |  |  | 0·520 |
| 0 | 453 (10·8%) | 358 (10·7%) | 95 (11·3%) |  |
| 1 | 705 (16·8%) | 569 (17·0%) | 136 (16·2%) |  |
| 2 | 727 (17·3%) | 580 (17·3%) | 147 (17·5%) |  |
| 3 | 703 (16·8%) | 549 (16·4%) | 154 (18·4%) |  |
| 4 | 589 (14·0%) | 472 (14·1%) | 117 (13· 9%) |  |
| 5 | 396 (9·4%) | 320 (9·5%) | 76 (9·1%) |  |
| 6 | 282 (6·7%) | 227 (6·8%) | 55 (6·6%) |  |
| 7 | 163 (3· 9%) | 132 (3· 9%) | 31 (3· 7%) |  |
| 8 | 102 (2· 4%) | 86 (2· 6%) | 16 (1·9%) |  |
| 9 | 41 (1·0%) | 32 (1·0%) | 9 (1·1%) |  |
| 10 | 17 (0·4%) | 17 (0·5%) | 0 (0·0%) |  |
| 11 | 8 (0·2%) | 6 (0·2%) | 2 (0·2%) |  |
| 12 | 3 (0·1%) | 3 (0·1%) | 0 (0·0%) |  |
| 13 | 3 (0·1%) | 3 (0·1%) | 0 (0·0%) |  |
| 14 | 2 (0·0%) | 2 (0·1%) | 0 (0·0%) |  |
| 15 | 1 (0·0%) | 0 (0·0%) | 1 (0·1%) |  |
| Rurality (n, %) |  |  |  | 0·555 |
| Rural | 1313 (31·3%) | 1058 (31·5%) | 255 (30·4%) |  |
| Urban | 2882 (68·7%) | 2298 (68·5%) | 584 (69·6%) |  |
| Gleason Score (n, %) |  |  |  | 0·820 |
| 4.0 | 29 (0·7%) | 24 (0·7%) | 5 (0·6%) |  |
| 5·0 | 46 (1·1%) | 38 (1·1%) | 8 (1·0%) |  |
| 5·5 | 23 (0·5%) | 18 (0·5%) | 5 (0·6%) |  |
| 6·0 | 456 (10·9%) | 355 (10·6%) | 101 (12· 0%) |  |
| 6·5 | 3 (0·1%) | 2 (0·1%) | 1 (0·1%) |  |
| 7·0 | 680 (16·2%) | 544 (16·2%) | 136 (16·2%) |  |
| 7·5 | 498 (11·9%) | 394 (11·7%) | 104 (12· 4%) |  |
| 8·0 | 802 (19·1%) | 657 (19·6%) | 145 (17·3%) |  |
| 8·5 | 29 (0·7%) | 23 (0·7%) | 6 (0·7%) |  |
| 9·0 | 995 (23·7%) | 782 (23·3%) | 213 (25·4%) |  |
| 9·5 | 369 (8·8%) | 302 (9·0%) | 67 (8·0%) |  |
| 10·0 | 265 (6·3%) | 217 (6·5%) | 48 (5·7%) |  |
| PSA at Diagnosis (median [Q1-Q3]) | 5·8 [0·8-22·0] | 5·8 [0·8-23·1] | 5·7 [0·6-18·4] | 0·081 |
| PSA at start of treatment (median [Q1-Q3]) | 31·7 [9·5-107·5] | 32·5 [9·9-110·1] | 30·0 [8·1-97·4] | 0·094 |
| g-rate (median [Q1-Q3]) | -5·4 [-6·9--4·6] | -5·4 [-7·0--4·6] | -5·3 [-6·7--4·6] | 0·390 |
| g-rate at 3months (median [Q1-Q3]) | -5·3 [-9·0--4·5] | -5·3 [-9·0--4·5] | -5·3 [-9·1--4·5] | 0·896 |
| g-rate at 6months (median [Q1-Q3]) | -5·4 [-7·1--4·6] | -5·4 [-7·1--4·6] | -5·3 [-6·8--4·6] | 0·367 |
| HGB at start of treatment (median [Q1-Q3]) | 12·0 [10·8-13·1] | 12·0 [10·8-13·1] | 12·2 [10·8-13·2] | 0·126 |
| WBC at start of treatment (median [Q1-Q3]) | 6·8 [5·4-8·5] | 6·8 [5·3-8·6] | 6·9 [5·4-8·4] | 0·655 |
| ANC at start of treatment (median [Q1-Q3]) | 4·5 [3·3-6·1] | 4·5 [3·3-6·1] | 4·5 [3·4-6·0] | 0·886 |
| PLT at start of treatment (median [Q1-Q3]) | 233·0 [188·0-287·0] | 234·0 [189·0-287·0] | 229·0 [185·5-287·5] | 0·313 |
| status (n, %) |  |  |  | 1·000 |
| 0·0 | 670 (16·0%) | 536 (16·0%) | 134 (16·0%) |  |
| 1·0 | 3525 (84·0%) | 2820 (84·0%) | 705 (84·0%) |  |
| time (median [Q1-Q3]) | 14·0 [7·2-25·5] | 13·9 [7·2-25·2] | 14·4 [7·3-26·2] | 0·447 |
| *p-values calculated using Chi-square test for categorical variables and Mann-Whitney U test for continuous variables.  Abbreviations: PSA, prostate-specific antigen (PSA); HGB, Hemoglobin; WBC, White Blood Cell Count; ANC, Absolute Neutrophil Count; PLT, Platelet. | | | | |

**Supplementary Table 3: Baseline Characteristics between training and test set in the third line of treatment**

|  | **Overall (n=1899)** | **Training (n=1519)** | **Test (n=380)** | **p_value** |
| --- | --- | --- | --- | --- |
| Age (median [Q1-Q3]) | 71·6 [67·0-76·4] | 71·6 [67·1-76·6] | 71·5 [66·7-75·7] | 0·453 |
| Race (n, %) |  |  |  | 0·773 |
| American Indian or Alaska Native | 11 (0·6%) | 10 (0·7%) | 1 (0·3%) |  |
| Asian | 10 (0·5%) | 9 (0·6%) | 1 (0·3%) |  |
| Black | 536 (28·2%) | 431 (28·4%) | 105 (27·6%) |  |
| Native Hawaiian or Other Pacific Islander | 13 (0·7%) | 11 (0·7%) | 2 (0·5%) |  |
| White | 1329 (70·0%) | 1058 (69·7%) | 271 (71·3%) |  |
| Charles Comorbidity Index without cancer (n, %) |  |  |  | 0·020 |
| 0 | 203 (10·7%) | 159 (10·5%) | 44 (11·6%) |  |
| 1 | 324 (17·1%) | 250 (16·5%) | 74 (19·5%) |  |
| 2 | 361 (19·0%) | 304 (20·0%) | 57 (15·0%) |  |
| 3 | 309 (16·3%) | 245 (16·1%) | 64 (16·8%) |  |
| 4 | 283 (14·9%) | 225 (14·8%) | 58 (15·3%) |  |
| 5 | 175 (9·2%) | 142 (9·3%) | 33 (8·7%) |  |
| 6 | 114 (6·0%) | 94 (6·2%) | 20 (5·3%) |  |
| 7 | 65 (3·4%) | 55 (3·6%) | 10 (2·6%) |  |
| 8 | 39 (2·1%) | 31 (2·0%) | 8 (2·1%) |  |
| 9 | 11 (0·6%) | 4 (0·3%) | 7 (1·8%) |  |
| 10 | 8 (0·4%) | 6 (0·4%) | 2 (0·5%) |  |
| 11 | 3 (0·2%) | 2 (0·1%) | 1 (0·3%) |  |
| 12 | 3 (0·2%) | 2 (0·1%) | 1 (0·3%) |  |
| 13 | 1 (0·1%) | 0 (0·0%) | 1 (0·3%) |  |
| Rurality (n, %) |  |  |  | 0·640 |
| Rural | 581 (30·6%) | 469 (30·9%) | 112 (29·5%) |  |
| Urban | 1318 (69·4%) | 1050 (69·1%) | 268 (70·5%) |  |
| Gleason Score (n, %) |  |  |  | 0·607 |
| 4·0 | 12 (0·6%) | 11 (0·7%) | 1 (0·3%) |  |
| 5·0 | 22 (1·2%) | 19 (1·3%) | 3 (0·8%) |  |
| 5·5 | 9 (0·5%) | 6 (0·4%) | 3 (0·8%) |  |
| 6·0 | 176 (9·3%) | 138 (9·1%) | 38 (10·0%) |  |
| 6·5 | 3 (0·2%) | 3 (0·2%) | 0 (0·0%) |  |
| 7·0 | 311 (16·4%) | 248 (16·3%) | 63 (16·6%) |  |
| 7·5 | 220 (11·6%) | 175 (11·5%) | 45 (11·8%) |  |
| 8·0 | 354 (18·6%) | 294 (19·4%) | 60 (15·8%) |  |
| 8·5 | 16 (0·8%) | 13 (0·9%) | 3 (0·8%) |  |
| 9·0 | 490 (25·8%) | 393 (25·9%) | 97 (25·5%) |  |
| 9·5 | 170 (9·0%) | 134 (8·8%) | 36 (9·5%) |  |
| 10·0 | 116 (6·1%) | 85 (5·6%) | 31 (8·2%) |  |
| PSA at Diagnosis (median [Q1-Q3]) | 5·5 [0·8-22·4] | 5·3 [0·8-22·6] | 6·1 [1·1-21·1] | 0·345 |
| PSA at start of treatment (median [Q1-Q3]) | 59·7 [17·6-175·4] | 59·0 [17·5-175·1] | 60·6 [18·1-185·1] | 0·429 |
| g-rate (median [Q1-Q3]) | -5·2 [-6·4--4·5] | -5·2 [-6·4--4·5] | -5·2 [-6·8--4·4] | 0·531 |
| g-rate at 3months (median [Q1-Q3]) | -5·1 [-8·9--4·4] | -5·1 [-8·9--4·4] | -5·0 [-8·9--4·4] | 0·945 |
| g-rate at 6months (median [Q1-Q3]) | -5·2 [-6·5--4·5] | -5·2 [-6·4--4·5] | -5·2 [-7·4--4·4] | 0·582 |
| HGB at start of treatment (median [Q1-Q3]) | 11·6 [10·4-12·8] | 11·7 [10·4-12·8] | 11·5 [10·4-12·7] | 0·276 |
| WBC at start of treatment (median [Q1-Q3]) | 6·8 [5·2-8·9] | 6·9 [5·2-8·9] | 6·7 [5·0-8·9] | 0·479 |
| ANC at start of treatment (median [Q1-Q3]) | 4·6 [3·2-6·4] | 4·6 [3·2-6·4] | 4·3 [3·2-6·4] | 0·417 |
| PLT at start of treatment (median [Q1-Q3]) | 239·0 [190·0-298·0] | 242·0 [192·0-299·0] | 235·0 [184·8-295·2] | 0·228 |
| status (n, %) |  |  |  | 1·000 |
| 0·0 | 261 (13·7%) | 209 (13·8%) | 52 (13·7%) |  |
| 1·0 | 1638 (86·3%) | 1310 (86·2%) | 328 (86·3%) |  |
| time (median [Q1-Q3]) | 11·4 [6·2-20·0] | 11·5 [6·2-20·1] | 11·0 [6·1-19·5] | 0·508 |
| *p-values calculated using Chi-square test for categorical variables and Mann-Whitney U test for continuous variables.  Abbreviations: PSA, prostate-specific antigen (PSA); HGB, Hemoglobin; WBC, White Blood Cell Count; ANC, Absolute Neutrophil Count; PLT, Platelet. | | | | |

**Supplementary Table 4: Baseline Characteristics between training and test set in the fourth line of treatment**

|  | **Overall (n=771)** | **Training (n=616)** | **Test (n=155)** | **p_value** |
| --- | --- | --- | --- | --- |
| Age (median [Q1-Q3]) | 71·4 [67·2-75·3] | 71·1 [67·1-75·2] | 72·2 [67·2-76·1] | 0·293 |
| Race (n, %) |  |  |  | 0·800 |
| American Indian or Alaska Native | 6 (0·8%) | 5 (0·8%) | 1 (0·6%) |  |
| Asian | 4 (0·5%) | 3 (0·5%) | 1 (0·6%) |  |
| Black | 233 (30·2%) | 185 (30·0%) | 48 (31·0%) |  |
| Native Hawaiian or Other Pacific Islander | 6 (0·8%) | 6 (1·0%) | 0 (0·0%) |  |
| White | 522 (67·7%) | 417 (67·7%) | 105 (67·7%) |  |
| Charles Comorbidity Index without cancer (n, %) |  |  |  | 0·529 |
| 0 | 79 (10·2%) | 68 (11·0%) | 11 (7·1%) |  |
| 1 | 150 (19·5%) | 123 (20·0%) | 27 (17·4%) |  |
| 2 | 153 (19·8%) | 114 (18·5%) | 39 (25·2%) |  |
| 3 | 121 (15·7%) | 92 (14·9%) | 29 (18·7%) |  |
| 4 | 104 (13·5%) | 86 (14·0%) | 18 (11·6%) |  |
| 5 | 72 (9·3%) | 59 (9·6%) | 13 (8·4%) |  |
| 6 | 36 (4·7%) | 26 (4·2%) | 10 (6·5%) |  |
| 7 | 31 (4·0%) | 25 (4·1%) | 6 (3·9%) |  |
| 8 | 15 (1·9%) | 14 (2·3%) | 1 (0·6%) |  |
| 9 | 3 (0·4%) | 2 (0·3%) | 1 (0·6%) |  |
| 10 | 2 (0·3%) | 2 (0·3%) | 0 (0·0%) |  |
| 11 | 2 (0·3%) | 2 (0·3%) | 0 (0·0%) |  |
| 12 | 2 (0·3%) | 2 (0·3%) | 0 (0·0%) |  |
| 14 | 1 (0·1%) | 1 (0·2%) | 0 (0·0%) |  |
| Rurality (n, %) |  |  |  | 0·591 |
| Rural | 230 (29·8%) | 187 (30·4%) | 43 (27·7%) |  |
| Urban | 541 (70·2%) | 429 (69·6%) | 112 (72·3%) |  |
| Gleason Score (n, %) |  |  |  | 0·219 |
| 4·0 | 2 (0·3%) | 2 (0·3%) | 0 (0·0%) |  |
| 5·0 | 6 (0·8%) | 4 (0·6%) | 2 (1·3%) |  |
| 5·5 | 1 (0·1%) | 1 (0·2%) | 0 (0·0%) |  |
| 6·0 | 70 (9·1%) | 52 (8·4%) | 18 (11·6%) |  |
| 7·0 | 124 (16·1%) | 97 (15·7%) | 27 (17·4%) |  |
| 7·5 | 83 (10·8%) | 63 (10·2%) | 20 (12·9%) |  |
| 8·0 | 160 (20·8%) | 141 (22·9%) | 19 (12·3%) |  |
| 8·5 | 4 (0·5%) | 4 (0·6%) | 0 (0·0%) |  |
| 9·0 | 213 (27·6%) | 166 (26·9%) | 47 (30·3%) |  |
| 9·5 | 64 (8·3%) | 53 (8·6%) | 11 (7·1%) |  |
| 10·0 | 44 (5·7%) | 33 (5·4%) | 11 (7·1%) |  |
| PSA at Diagnosis (median [Q1-Q3]) | 6·0 [0·8-24·6] | 5·9 [0·9-24·6] | 6·3 [0·6-26·3] | 0·679 |
| PSA at start of treatment (median [Q1-Q3]) | 97·1 [33·8-282·4] | 101·6 [34·6-316·9] | 80·6 [27·7-185·4] | 0·047 |
| g-rate (median [Q1-Q3]) | -4·9 [-5·8--4·4] | -5·0 [-5·8--4·4] | -4·9 [-5·8--4·4] | 0·804 |
| g-rate at 3months (median [Q1-Q3]) | -4·9 [-5·9--4·3] | -4·9 [-6·0--4·3] | -4·8 [-5·8--4·4] | 0·693 |
| g-rate at 6months (median [Q1-Q3]) | -4·9 [-5·8--4·4] | -5·0 [-5·8--4·4] | -4·9 [-5·8--4·4] | 0·805 |
| HGB at start of treatment (median [Q1-Q3]) | 11·2 [9·8-12·2] | 11·1 [9·8-12·2] | 11·3 [9·6-12·2] | 0·982 |
| WBC at start of treatment (median [Q1-Q3]) | 6·7 [4·9-9·1] | 6·7 [4·9-9·0] | 6·8 [5·1-9·3] | 0·632 |
| ANC at start of treatment (median [Q1-Q3]) | 4·6 [3·2-6·5] | 4·6 [3·2-6·5] | 4·7 [3·1-6·7] | 0·677 |
| PLT at start of treatment (median [Q1-Q3]) | 238·0 [188·0-294·0] | 239·0 [188·0-294·0] | 231·0 [187·5-296·0] | 0·714 |
| status (n, %) |  |  |  | 1·000 |
| 0·0 | 103 (13·4%) | 82 (13·3%) | 21 (13·5%) |  |
| 1·0 | 668 (86·6%) | 534 (86·7%) | 134 (86·5%) |  |
| time (median [Q1-Q3]) | 8·6 [4·9-15·1] | 8·7 [4·7-15·7] | 8·5 [5·5-14·2] | 0·994 |
| *p-values calculated using Chi-square test for categorical variables and Mann-Whitney U test for continuous variables.  Abbreviations: PSA, prostate-specific antigen (PSA); HGB, Hemoglobin; WBC, White Blood Cell Count; ANC, Absolute Neutrophil Count; PLT, Platelet. | | | | |

|  | **Censorship**  **(%)** | **Feature set** | **G3Surv** | | | | **G6Surv** | | | | **GfSurv** | | | |
| --- | --- | --- | --- | --- | --- | --- | --- | --- | --- | --- | --- | --- | --- | --- |
|  |  |  | C-index  (Harrel) | C-index  (Uno) | IBS | tAUC | C-index  (Harrell) | C-index  (Uno) | IBS | tAUC | C-index  (Harrell) | C-index  (Uno) | IBS | tAUC |
| **First treatment line** | 23·5 | 10 features | 0·746 (0·733-0·759) | 0·729 (0·716-0·742) | 0·077 (0·068 - 0·087) | 0·822 (0·806 - 0·839) | 0·754 (0·741-0·766) | 0·736 (0·723-0·749) | 0·076 (0·067 - 0·087) | 0·831 (0·815 - 0·847) | 0·761 (0·748-0·775) | 0·745 (0·732-0·759) | 0·074 (0·066 - 0·085) | 0·836 (0·820 - 0·852) |
|  |  | 7 features | 0·744 (0·731-0·757) | 0·726 (0·713-0·739) | 0·077 (0·069 - 0·088) | 0·821 (0·805 - 0·837) | 0·751 (0·738-0·764) | 0·733 (0·720-0·746) | 0·076 (0·067 - 0·087) | 0·829 (0·813 - 0·845) | 0·759 (0·746-0·772) | 0·742 (0·730-0·756) | 0·074 (0·065 - 0·085) | 0·835 (0·818 - 0·851) |
| **Second treatment line** | 16.0 | 10 features | 0·737 (0·719-0·755) | 0·726 (0·710-0·744) | 0·091 (0·082 - 0·102) | 0·818 (0·797 - 0·840) | 0·743 (0·726-0·761) | 0·733 (0·716-0·750) | 0·088 (0·079 - 0·100) | 0·825 (0·804 - 0·845) | 0·754 (0·737-0·772) | 0·744 (0·727-0·761) | 0·085 (0·076 - 0·095) | 0·835 (0·814 - 0·855) |
|  |  | 7 features | 0·736 (0·718-0·754) | 0·725 (0·708-0·743) | 0·091 (0·082 - 0·102) | 0·818 (0·796 - 0·840) | 0·742 (0·725-0·760) | 0·732 (0·716-0·749) | 0·089 (0·079 - 0·099) | 0·823 (0·802 - 0·844) | 0·753 (0·736-0·770) | 0·743 (0·727-0·760) | 0·085 (0·076 - 0·095) | 0·833 (0·812 - 0·853) |
| **Third treatment line** | 13· 7 | 10 features | 0·704 (0·675-0·731) | 0·697 (0·668-0·725) | 0·076 (0·063 - 0·097) | 0·786 (0·750 - 0·819) | 0·717 (0·688-0·745) | 0·709 (0·680-0·737) | 0·076 (0·064 - 0·098) | 0·801 (0·765 - 0·835) | 0·724 (0·696-0·750) | 0·716 (0·688-0·743) | 0·075 (0·063 - 0·096) | 0·809 (0·774 - 0·842) |
|  |  | 7 features | 0·705 (0·679-0·732) | 0·698 (0·671-0·724) | 0·077 (0·064 - 0·099) | 0·789 (0·755 - 0·822) | 0·720 (0·692-0·748) | 0·712 0·684-0·739) | 0·076 (0·063 - 0·098) | 0·805 (0·768 - 0·838) | 0·730 (0·703-0·756) | 0·722 (0·695-0·748) | 0·075 (0·063 - 0·096) | 0·816 (0·781 - 0·848) |
| **Fourth treatment line** | 13·6 | 10 features | 0·700 (0·647-0·750) | 0·694 (0·643-0·742) | 0·096 (0·077 - 0·120) | 0·766 (0·700 - 0·830) | 0·698 (0·649-0·745) | 0·694 (0·646-0·739) | 0·094 (0·076 - 0·118) | 0·763 (0·696 - 0·825) | 0·701 (0·651-0·749) | 0·697 (0·649-0·744) | 0·093 (0·075 - 0·117) | 0·766 (0·700 - 0·828) |
|  |  | 7 features | 0·695 (0·645-0·744) | 0·689 (0·641-0·736) | 0·096 (0·078 - 0·121) | 0·763 (0·695 - 0·826) | 0·700 (0·652-0·747) | 0·695 (0·648-0·741) | 0·093 (0·076 - 0·117) | 0·765 (0·700 - 0·825) | 0·702 (0·653-0·749) | 0·697 (0·649-0·745) | 0·093 (0·076 - 0·117) | 0·768 (0·702 - 0·830) |

**Supplementary Table 5: Comparison of performance metrics for GxSurv using 10 features versus 7 features for treatment outcome in mCRPC.** The 7-feature model excludes PSA at diagnosis, White Blood Cell Count, and Charlson Comorbidity Index without cancer.

Supplementary Table 6:Comparison of performance metrics between GxSurv and CoxPH models using all features.

|  | **Censorship**  **(%)** | **Models** | **3-month g-rateurv** | | | | **6-month g-rate** | | | | **final g-rate** | | | |
| --- | --- | --- | --- | --- | --- | --- | --- | --- | --- | --- | --- | --- | --- | --- |
|  |  |  | C-index  (Harrel) | C-index  (Uno) | IBS | tAUC | C-index  (Harrell) | C-index  (Uno) | IBS | tAUC | C-index  (Harrell) | C-index  (Uno) | IBS | tAUC |
| **First treatment line** | 23·5 | GxSurv | 0·746 (0·733-0·759) | 0·729 (0·716-0·742) | 0·077 (0·068 - 0·087) | 0·822 (0·806 - 0·839) | 0·754 (0·741-0·766) | 0·736 (0·723-0·749) | 0·076 (0·067 - 0·087) | 0·831 (0·815 - 0·847) | 0·761 (0·748-0·775) | 0·745 (0·732-0·759) | 0·074 (0·066 - 0·085) | 0·836 (0·820 - 0·852) |
|  |  | CoxPH | 0·710 (0·694- 0·724) | 0·693  (0·678- 0·706) | 0·163  (0·157- 0·169) | 0·783  (0·764- 0·801) | 0·710  (0·696- 0·725) | 0·695  (0·681- 0·708) | 0·161  (0·156- 0·167) | 0·782  (0·763, 0·799) | 0·718 (0·703- 0·732) | 0·704 (0·690- 0·717) | 0·157 (0·151- 0·163) | 0·787 (0·766- 0·805) |
| **Second treatment line** | 16.0 | GxSurv | 0·737 (0·719-0·755) | 0·726 (0·710-0·744) | 0·091 (0·082 - 0·102) | 0·818 (0·797 - 0·840) | 0·743 (0·726-0·761) | 0·733 (0·716-0·750) | 0·088 (0·079 - 0·100) | 0·825 (0·804 - 0·845) | 0·754 (0·737-0·772) | 0·744 (0·727-0·761) | 0·085 (0·076 - 0·095) | 0·835 (0·814 - 0·855) |
|  |  | CoxPH | 0·703 (0·683- 0·723) | 0·694  (0·675- 0·713) | 0·168 (0·160- 0·175) | 0·783  (0·760- 0·809) | 0·708  (0·689- 0·728) | 0·700  (0·682- 0·719) | 0·167  (0·159- 0·175) | 0·782 (0·757, 0·807) | 0·712 (0·691- 0·733) | 0·705 (0·686- 0·725) | 0·166 (0·157- 0·174) | 0·784  (0·756- 0·811) |
| **Third treatment line** | 13· 7 | GxSurv | 0·704 (0·675-0·731) | 0·697 (0·668-0·725) | 0·076 (0·063 - 0·097) | 0·786 (0·750 - 0·819) | 0·717 (0·688-0·745) | 0·709 (0·680-0·737) | 0·076 (0·064 - 0·098) | 0·801 (0·765 - 0·835) | 0·724 (0·696-0·750) | 0·716 (0·688-0·743) | 0·075 (0·063 - 0·096) | 0·809 (0·774 - 0·842) |
|  |  | CoxPH | 0·697 (0·668- 0·724) | 0·689  (0·660- 0·717) | 0·169 (0·159- 0·180) | 0·777  (0·740- 0·809) | 0·695  (0·665- 0·721) | 0·687  (0·658- 0·713) | 0·170  (0·159- 0·181) | 0·771 (0·730, 0·806) | 0·700 (0·669- 0·730) | 0·694 (0·664- 0·723) | 0·168 (0·157- 0·179) | 0·771  (0·728- 0·808) |
| **Fourth treatment line** | 13·6 | GxSurv | 0·700 (0·647-0·750) | 0·694 (0·643-0·742) | 0·096 (0·077 - 0·120) | 0·766 (0·700 - 0·830) | 0·698 (0·649-0·745) | 0·694 (0·646-0·739) | 0·094 (0·076 - 0·118) | 0·763 (0·696 - 0·825) | 0·701 (0·651-0·749) | 0·697 (0·649-0·744) | 0·093 (0·075 - 0·117) | 0·766 (0·700 - 0·828) |
|  |  | CoxPH | 0·675 (0·615- 0·731) | 0·669  (0·611- 0·725) | 0·177 (0·158- 0·198) | 0·732  (0·652- 0·805) | 0·690  (0·635- 0·746) | 0·685  (0·629- 0·740) | 0·172  (0·154- 0·192) | 0·750 (0·674, 0·823) | 0·693 (0·634- 0·749) | 0·688 (0·628- 0·743) | 0·171 (0·152- 0·193) | 0·753 (0·675- 0·826) |

**Supplementary Table 7: Comparison of predictive performance of G0Surv and CoxPH without inclusion of g-rate using all other features for treatment outcome.**

|  | **Censorship(%)** | **Models** | **Without g-rate** | | | |
| --- | --- | --- | --- | --- | --- | --- |
|  |  |  | **C-index**  **(Harrell)** | **C-index**  **(Uno)** | **IBS** | **tAUC** |
| First treatment line | 23·5 | G0Surv | 0·712 (0·697-0·726) | 0·698 (0·684-0·711) | 0·081 (0·071 - 0·092) | 0·780 (0·762 - 0·798) |
|  |  | CoxPH | 0·664 (0·648-0·680) | 0·652 (0·637- 0·667) | 0·177 (0·172-0·182) | 0·725 (0·703-0·746) |
| Second treatment line | 16.0 | G0Surv | 0·688 (0·667-0·706) | 0·678 (0·658-0·696) | 0·100 (0·090 - 0·112) | 0·760 (0·732 - 0·784) |
|  |  | CoxPH | 0·665 (0·643-0·689) | 0·656  (0·635 - 0·679) | 0·183 (0·176 -0·190) | 0·734 (0·705-0·765) |
| Third treatment line | 13·7 | G0Surv | 0·647 (0·612-0·680) | 0·643 (0·607-0·675) | 0·083 (0·070 - 0·106) | 0·707 (0·660 - 0·751) |
|  |  | CoxPH | 0·662 (0·631- 0·693) | 0·656 (0·626 - 0·686) | 0·179 (0·169- 0·188) | 0·731 (0·689 - 0·770) |
| Fourth treatment line | 13·6 | G0Surv | 0·625 (0·569-0·680) | 0·621 (0·566-0·676) | 0·105 (0·085 - 0·132) | 0·664 (0·590 - 0·741) |
|  |  | CoxPH | 0·595 (0·534-0·655) | 0·589  (0·520-0·646) | 0·195 (0·177- 0·214) | 0·635 (0·550-0·716) |

**Supplementary Table 8: Comparison of treatment outcome performance between previous studies and the proposed G3Surv model across first- to fourth-line treatments.** Model performance is evaluated using Harrell’s C-index, Uno’s C-index, Integrated Brier Score (IBS), and time-dependent area under the curve (tAUC), with values presented alongside 95% confidence interval. In the overall cohort, G3Surv demonstrated higher predictive performance than the model reported by Moreira et al. -: Not available. Bold represents the best performance.

| Treatment line | Metrics | Halabi et al.(^21^) | Halabi et al.(^22^) | Moreira et al.(^45^) | G3Surv (Ours) |
| --- | --- | --- | --- | --- | --- |
| First-line mCRPC | Harrell’s C-index | – | – |  | 0·746 (95% CI, 0·733 to 0·759) |
|  | Uno’s C-index | – | – |  | 0·729 (95% CI, 0·716 to 0·742) |
|  | IBS | – | – |  | 0·077 (95% CI, 0·068 to 0·087) |
|  | tAUC | 0·73 (95% CI, 0·70 to 0·73) | – |  | **0·822 (95% CI, 0·806 to 0·839)** |
| Second-line mCRPC | Harrell’s C-index | – | – |  | 0·737 (95% CI, 0·719 to 0·755) |
|  | Uno’s C-index | – | – |  | 0·726 (95% CI, 0·710 to 0·744) |
|  | IBS | – | – |  | 0·091 (95% CI, 0·082 to 0·102) |
|  | tAUC | – | 0·73 (95% CI, 0·72 to 0·74) |  | **0·818 (95% CI, 0·797 to 0·840)** |
| Third-line mCRPC | Harrell’s C-index | – | – |  | 0·704 (95% CI, 0·675 to 0·731) |
|  | Uno’s C-index | – | – |  | 0·697 (95% CI, 0·668 to 0·725) |
|  | IBS | – | – |  | 0·076 (95% CI, 0·063 to 0·097) |
|  | tAUC | – | – |  | **0·786 (95% CI, 0·750 to 0·819)** |
| Fourth-line mCRPC | Harrell’s C-index | – | – |  | 0·700 (95% CI, 0·647 to 0·750) |
|  | Uno’s C-index | – | – |  | 0·694 (95% CI, 0·643 to 0·742) |
|  | IBS | – | – |  | 0·096 (95% CI, 0·077 to 0·120) |
|  | tAUC | – | – |  | **0·766 (95% CI, 0·700 to 0·830)** |
| Overall mCRPC | Harrell’s C-index | – | – | 0·67(CI not reported) | **0·755 (95% CI, 0·745 to 0·764)** |
|  | Uno’s C-index | – | – | – | 0·742 (95% CI, 0·733 to 0·751) |
|  | IBS | – | – | – | 0·062 (95% CI, 0·056 to 0·069) |
|  | tAUC | – | – | – | 0·835 (95% CI, 0·823 to 0·845) |
